## Supplement for "Assessing the impact of SARS-CoV-2 prevention measures in Austrian schools by means of agent-based simulations calibrated to cluster tracing data" (from another version of this preprint: version 1, 2021-04-19)

#### Supplementary Note 1: School types

Below we describe the four different school types included in our model: (i) primary schools, (ii) lower secondary schools, (iii) upper secondary schools, and (iv) secondary schools. These four school types accommodate 955 576 out of a total of 1 113 673 or 85.8% of students in Austria, as of the Austrian school statistic 2017/18 (1). The school types *not* included in our model are characterized by their small number of schools per type and complexity of organisation. These school types include for example specialized sports schools, private schools and foreign schools.

**A. Primary schools.** Austrian primary schools are compulsory for children above the age of 6 and offer education according to ISCED level 1. Austrian primary schools have 4 levels, therefore students in Austrian primary schools are typically aged between 6 and 10. According to the Austrian education statistics of the year 2017/18 (1), there are 3033 Primary schools with a total of 18,245 classes and 339,382 students in Austria. Therefore the average Austrian primary school has 8 classes with 19 students each, corresponding to two classes per level.

About 50% of primary schools in Austria offer full-day-care and about 25% of students in primary schools participate in full-day-care (2). Full-day-care is organised in two different models: integrated and separated. In the separated model, teaching ends after 4-5 hours every day and is followed by supervised lunch, work on homework and other activities for another 3-4 hours. Since usually not all students of a school participate in the full-day-care in the separated model, classes are broken up after the teaching part of the day and students are re-distributed into a number of groups that is usually smaller than the number of classes the school has. In the integrated model, teaching is intertwined with other activities and supervised work on homework. In this model, all students participate in the full-day-care and classes are not broken up. Of the schools that offer full-day-care, 95% run the separated model (3). To approximate these numbers, we assume that 50% of schools in our primary school ensemble offer full-day-care in which 50% of the children in these schools participate, respectively. We only model separated full-day-care, since this corresponds to the vast majority of Austrian primary schools. For afternoon supervision, students are re-organised into groups of the same size as the morning classes by randomly selecting 50% of the total number of students and randomly assigning them to M groups, where M=N/2 and N is the number of classes (4 in the case of the average primary school).

According to the syllabus (4) for Austrian primary schools, Austrian kids spend between 20 and 25 hours at school every week. Out of this total amount of hours, 11 hours are spent learning primary subjects (reading, writing, math, information retrieval), the other hours are spent learning minor subjects (sports, religion, creative subjects). We approximate this syllabus in the following way: Children in our simulated primary schools spend 20 hours / week or 4 hours / day at school. Every day, the first two hours are allotted to learning primary subjects, whereas the rest of the day is alloted to secondary subjects. Primary subjects are all taught by a single teacher for every class, respectively ("class teacher"), whereas secondary subjects are taught by different teachers.

According to the financing regulations for Austrian primary schools (2), 1.5 teachers per class are financed in Austrian schools. Nevertheless, this number does not include staff needed for additional offers such as full-day-care, language classes or religion classes. According to the Austrian school statistics 2017/18, the total number of teachers in primary schools was 33050, whereas the number of classes was (1) 18245, yielding an average of 1.81 teachers per class. Unfortunately, this number cannot be stratified for schools with and without full-day-care. This number also only reflects the number of full-time equivalents alloted for teaching and not the true number of persons that are involved in teaching, which might be higher if a share of teachers only works part-time. According to the structured interview we conducted with a director of a primary school offering full-day-care, the number of teachers was 14 for a school with 6 classes, yielding an average of 2.3 teachers per class. Since the primary school we interviewed offered full-day-care organised into 4 afternoon supervision groups, this number included personnel to supervise students in the afternoon.

To approximate this situation, we model schools without full-day-care to have 1.5 teachers per class, and each teacher has contact to a total of two different classes (every day, but also over the course of the week). The low number of classes reflects the fact that teaching in primary schools is still largely organized around a focal class teacher that teaches the same class for extended periods of time in several primary subjects. We model schools with full-day-care to have 2 teachers per class, where each teacher still has contact to a total of two different classes, and half of the teachers supervise an afternoon care group in addition to teaching. This is also consistent with the information we received in our interview of a primary school director, about the number of different classes a teacher sees.

A visualization of the contact network of an average Austrian primary school with daycare is schown in the main manuscript, Figure **2. A**.

**B. Lower secondary schools.** Lower secondary schools offer education according to ISCED level 2 and teach children aged 11 to 14. In Austria, there are two main school types that offer this type of education: (i) "Sonderschule" (special needs school) with 292 schools, 1626 classes and 14815 students (1, 5) and (ii) "Mittelschule" (middle school) with 1178 schools, 10458 classes and 207943 students (1, 6). We note that for the type "Mittelschule" the numbers that are reported in the latest statistics from the year 2017/18 for the types "Hauptschule" (47 schools) and "Neue Mittelschule" (1131 schools) were combined, since the Hauptschule was phased out by September 2020 and replaced by the Mittelschule. Another school type that offers ISCED level 2 education in Austria is the "AHS Unterstufe" (281 schools, 4901 classes, 119591 students), which usually is combined with an "AHS Oberstufe" that offers ISCED level 3 education. These combined schools are modelled separately and described below in Section D. We therefore model the average lower secondary school as having $(1626 + 10458)/(292 + 1178) = 8$ classes with $(14815 + 207943)/(1626 + 10458) = 18$ students per class.

Similar to primary schools, in these school types about 50% of schools offer daycare (2) for children in the afternoons: in the Mittelschule 18% of the children participate in daycare and in the Sonderschule 28% of the children participate in daycare. The average weighted by the number of classes in each school type results in 19% of children participating in daycare. Therefore in the schools that do offer daycare, on average 38% of the students participate in daycare (a bit less than in primary schools, where the ratio is 50%). Similar to primary schools, the vast majority of these schools run the separated model and we model the daycare similarly. We therefore model these schools similarly to schools offering primary education when it comes to daycare: We construct an ensemble of schools where 50% of schools offer daycare. In the schools that do offer daycare, 38% of students are randomly selected and assigned to M daycare supervision groups, where $M = N/2$ and $N$ is the number of classes the school has (4 in the case of the average lower secondary school).

According to the syllabus, students in the Mittelschule spend an average of 30 hours per week, or 6 hours per day at school (7). The syllabus includes on average 12 different subjects per school year (some subjects are not taught in all years). According to the syllabus of the Sonderschule, children spend a bit less time (on average 28 hours per week) in these schools (8), the number of subjects is the same as in the other lower secondary school types.

The number of teachers per class in lower secondary schools is higher than in primary schools, since there are more subjects and no dedicated class teachers anymore (1). Sonderschulen have the highest number of teachers per class (3.4) followed by Mittelschulen (2.8). The average number of teachers per class in lower secondary education (weighted by the number of classes) is 2.87. Among the schools we interviewed, there were two MS, one with 15 classes, 36 teachers (excluding daycare personnel) and 2.4 teachers per class, and one with 16 classes, 50 teachers (including daycare personnel) and 3.1 teachers per class. Both of these schools offered daycare. Unfortunately, we cannot differentiate between schools that offer daycare and schools that do not in the Austria-wide statistics for the number of teachers per class in lower secondary schools. Given the average number of teachers per class for all lower secondary schools (with and without daycare) and our interviews, we think it is warranted to assume lower secondary schools with daycare employ 3 teachers per class, whereas schools without daycare employ 2.5 teachers per class.

For the school type Mittelschule, team-teaching has become the norm in Austria. This means that for the majority of classes (according to our interviews 4/6 to 5/6 of lessons), two teachers are present during a lesson. This obviously introduces additional contacts between the teachers that teach lessons together. In Sonderschulen it is also customary to have additional supporting personnel present to support students with special needs. We therefore model schools that teach lower secondary levels to have a large share of team-teaching: for schools without daycare, 4 out of 6 lessons every day are team-taught lessons. For schools with daycare, 5 out of 6 lessons every day are team-taught lessons.

**C. Upper secondary schools.** Upper secondary schools offer education according to ISCED level 3 and teach children aged 15 to 18. In Austria, there are a two main types of schools that offer ISCED level 3 education: (i) "Oberstufenrealgymnasium" (upper secondary school) with 114 schools, 1183 classes and 26211 students (1, 9) and (ii) "Berufsbildende mittlere & höhere Schule" (vocational school) with 734 schools, 8042 classes and 187592 students (1, 10). There is also the "AHS Oberstufe", which offers ISCED level 3 education in combination with the AHS Unterstufe (level 2 education) which is modelled separately and described below in Section D. In addition, there are the "polytechnischen Schulen", which only offer education for the 9th grade (students aged 15). There are a total of 237 polytechnische Schulen with 764 classes and 15309 students. Compared to the other two school types, only a minor share of students aged 15-18 attend these schools (7%). We therefore chose to not model this school type separately.

Following the school types that we want to model for upper secondary education, the average school offering upper secondary education has $(1183 + 8042)/(114 + 734) = 10$ classes (rounded down from 10.9 to achieve an even number of classes, needed for modeling), and $(26211 + 187592)/(1183 + 8042) = 23$ students per class.

None of the upper secondary education school types offer any daycare in the afternoon after lessons end.

Oberstufenrealgymnasien have the same syllabus as the AHS Oberstufe and are in general similar to the AHS Oberstufe. The only difference is that these schools do not offer lower secondary education. Nevertheless, we choose to model the Oberstufenrealgymnasium separately from the Allgemeinbildende höhere Schulen (AHS Unterstufe + AHS Oberstufe) since we think that the different age structure of the student population in these schools (only students aged 15-18 instead of students aged 11-18) warrants a different treatment. According to the syllabus of the AHS Oberstufe (11), students spend an average of 32.5 hours of lessons per week at school. In addition, there are a number of non-mandatory tutorials, longer breaks and vocational activities that students engage in. In summary, we model the students to spend 8 hours per day at school.

Unfortunately, the Austria-wide school statistics for the year 2017/18 (1) does not show the number of teachers per class stratified for the Oberstufenrealgymnasium. Since these schools are similar to the AHS Oberstufe, we use the number of teachers per class for the AHS Obserstufe, which have on average 2.5 teachers per class. The berufsbildende mittlere & höhere Schulen have on average 2.9 teachers per class. The average weighted by the number of classes of each school type is 2.85 teachers per class. Therefore we model this school type as using 2.85 teachers per class.

Teachers in these schools teach a different number of different classes depending on their subjects: teachers of major subjects (German language, math, first foreign language) usually see fewer classes for more hours (2-3 different classes per day according to our interviews). Teachers of minor subjects (Physics, Geography, Religion, etc.) usually see more classes for fewer hours (4-5 different classes per day according to our interviews). Teachers that teach a combination of major and minor subjects fall in-between this range and see 3-4 different classes per day according to our interviews. Also according to our interviews, the teacher population of a school falls within these three categories (only major, only minor, major + minor) to approximately equal parts. We therefore model the teacher population such that approximately 1/3 of teachers sees 2 classes per day, 1/3 of teachers sees 3 teachers per day and 1/3 of teachers sees 4-5 classes per day (depending on the number of classes the school has and the corresponding integer number of teachers). According to our interviews, there is a small amount of team-teaching in these schools, predominantly for language classes, in which a native speaking teacher supports the main teacher. Accordingly, approximately 10% of lessons taught in these schools are modelled as team-teaching lessons and contacts between teachers and other teachers and teachers and students are set accordingly.

**D. Secondary schools.** Next to schools that offer only lower secondary education or upper secondary education, there are schools that offer combined lower and upper secondary education (ISCED level 2 and 3) and teach children aged 11 to 18. There are 281 of these schools ("AHS Langform"), with 7610 classes and 179633 students (1).

Some of these schools offer daycare, but we choose to not include a separate model for these schools, since only a fraction of students (those aged 10-14) do attend daycare if it is offered and only a smaller fraction of time is covered by daycare, since students have a higher number of regular lessons per week as compared to primary schools.

According to the syllabus of the AHS Oberstufe (11), in these schools students have an average of 32.5 hours per week of lessons at school. In addition, there are a number of non-mandatory tutorials, longer breaks and vocational activities that students engage in. In summary, we model the students to spend 8 hours per day at school.

The average number of teachers per class for these schools is 2.5. Given the rather low number of teachers available (as compared to 3 teachers per class for schools with only upper secondary education), we do not model lessons with team teaching in these schools.

A visualization of the contact network of an average Austrian secondary school is shown in the main manuscript Figure **2. B**.

#### Supplementary Note 2: Tables for Main Results

| type | measure | cluster size, mean (75$^{th}$,90$^{th}$ percentile) student | cluster size, mean (75$^{th}$,90$^{th}$ percentile) teacher | $R$, mean (SD) student | $R$, mean (SD) teacher |
|---|---|---|---|---|---|
| primary | none | 8 (20, 36) | 12 (28, 42) | 1.8 (1.7) | 3.4 (3.0) |
| | ventilation | 2 (4, 6) | 3 (6, 10) | 0.9 (0.9) | 1.3 (1.5) |
| | masks teacher | 5 (14, 22) | 6 (14, 24) | 1.7 (1.6) | 1.7 (1.8) |
| | masks students | 2 (4, 8) | 5 (10, 18) | 0.9 (1.0) | 2.5 (2.4) |
| | class size reduction | 2 (4, 6) | 4 (8, 12) | 0.8 (1.0) | 1.9 (1.9) |
| | testing teacher 1x | 7 (16, 30) | 7 (18, 34) | 1.7 (1.7) | 2.4 (3.0) |
| | testing students 1x | 4 (8, 20) | 8 (20, 32) | 1.0 (1.3) | 3.1 (2.7) |
| | testing teacher 2x | 6 (14, 24) | 6 (14, 30) | 1.7 (1.7) | 2.0 (2.7) |
| | testing students 2x | 3 (4, 12) | 8 (20, 34) | 0.8 (1.2) | 3.4 (3.0) |
| + day care | none | 23 (73, 116) | 27 (82, 124) | 2.2 (2.0) | 3.2 (3.0) |
| | ventilation | 3 (6, 10) | 4 (6, 14) | 1.0 (1.2) | 1.3 (1.5) |
| | masks teacher | 15 (42, 78) | 14 (36, 76) | 2.1 (2.0) | 1.6 (1.8) |
| | masks students | 4 (6, 14) | 7 (16, 28) | 1.1 (1.2) | 2.4 (2.4) |
| | class size reduction | 3 (4, 10) | 5 (8, 14) | 0.8 (1.1) | 1.9 (2.1) |
| | testing teacher 1x | 21 (63, 114) | 17 (48, 96) | 2.2 (2.2) | 2.2 (2.9) |
| | testing students 1x | 8 (12, 46) | 16 (39, 78) | 1.3 (1.7) | 3.1 (2.9) |
| | testing teacher 2x | 20 (50, 120) | 14 (24, 90) | 2.2 (2.0) | 1.8 (2.6) |
| | testing students 2x | 7 (8, 32) | 16 (38, 80) | 1.1 (1.6) | 3.3 (2.9) |
| lower secondary | none | 57 (172, 192) | 66 (174, 196) | 2.6 (2.1) | 6.5 (5.5) |
| | ventilation | 5 (8, 22) | 8 (16, 34) | 1.2 (1.2) | 2.6 (2.5) |
| | masks teacher | 24 (65, 108) | 24 (65, 108) | 2.4 (2.1) | 3.3 (3.1) |
| | masks students | 13 (26, 76) | 22 (56, 96) | 1.3 (1.3) | 4.8 (4.2) |
| | class size reduction | 9 (16, 46) | 17 (38, 68) | 1.1 (1.2) | 3.8 (3.5) |
| | testing teacher 1x | 43 (137, 172) | 39 (138, 170) | 2.6 (2.2) | 4.4 (5.4) |
| | testing students 1x | 28 (92, 164) | 51 (154, 184) | 1.5 (1.8) | 6.2 (5.2) |
| | testing teacher 2x | 41 (126, 168) | 30 (100, 160) | 2.6 (2.1) | 3.5 (4.8) |
| | testing students 2x | 18 (15, 136) | 49 (145, 178) | 1.1 (1.6) | 6.5 (5.5) |
| + day care | none | 49 (148, 176) | 55 (154, 178) | 2.7 (2.3) | 4.3 (3.8) |
| | ventilation | 5 (8, 18) | 5 (10, 22) | 1.3 (1.3) | 1.8 (1.9) |
| | masks teacher | 25 (74, 116) | 22 (61, 110) | 2.6 (2.2) | 2.2 (2.3) |
| | masks students | 8 (16, 34) | 16 (34, 64) | 1.4 (1.4) | 3.4 (3.2) |
| | class size reduction | 5 (8, 20) | 10 (18, 36) | 1.1 (1.3) | 2.7 (2.7) |
| | testing teacher 1x | 41 (136, 166) | 34 (115, 162) | 2.6 (2.3) | 3.0 (3.8) |
| | testing students 1x | 22 (53, 130) | 36 (115, 152) | 1.5 (2.0) | 4.1 (3.7) |
| | testing teacher 2x | 36 (117, 152) | 27 (87, 142) | 2.6 (2.3) | 2.3 (3.2) |
| | testing students 2x | 13 (10, 90) | 31 (92, 140) | 1.2 (1.7) | 4.2 (3.8) |
| upper secondary | none | 113 (302, 330) | 122 (306, 332) | 3.4 (2.6) | 7.1 (6.0) |
| | ventilation | 8 (16, 34) | 11 (24, 52) | 1.6 (1.5) | 2.8 (2.6) |
| | masks teacher | 56 (166, 224) | 57 (171, 226) | 3.1 (2.4) | 3.6 (3.3) |
| | masks students | 26 (76, 136) | 36 (102, 150) | 1.7 (1.5) | 5.4 (4.6) |
| | class size reduction | 13 (18, 72) | 25 (60, 100) | 1.4 (1.3) | 4.1 (3.6) |
| | testing teacher 1x | 94 (276, 310) | 73 (264, 306) | 3.3 (2.5) | 5.0 (6.1) |
| | testing students 1x | 49 (201, 280) | 91 (260, 296) | 2.0 (2.4) | 7.4 (6.3) |
| | testing teacher 2x | 83 (268, 306) | 61 (236, 292) | 3.2 (2.4) | 3.9 (5.2) |
| | testing students 2x | 36 (45, 256) | 85 (260, 298) | 1.4 (2.0) | 7.3 (6.1) |
| secondary | none | 282 (746, 806) | 306 (760, 826) | 2.9 (2.4) | 8.6 (7.9) |
| | ventilation | 9 (13, 42) | 16 (32, 66) | 1.3 (1.3) | 3.3 (3.3) |
| | masks teacher | 109 (344, 478) | 111 (361, 486) | 2.7 (2.2) | 4.1 (4.0) |
| | masks students | 60 (180, 360) | 120 (344, 416) | 1.4 (1.3) | 6.7 (6.2) |
| | class size reduction | 34 (22, 220) | 81 (220, 278) | 1.3 (1.4) | 5.4 (5.1) |
| | testing teacher 1x | 208 (662, 732) | 190 (642, 738) | 2.8 (2.3) | 5.8 (7.1) |
| | testing students 1x | 128 (575, 710) | 261 (686, 748) | 1.7 (2.2) | 8.2 (7.6) |
| | testing teacher 2x | 188 (638, 710) | 161 (596, 702) | 2.8 (2.3) | 4.7 (6.3) |
| | testing students 2x | 107 (484, 680) | 248 (659, 736) | 1.3 (1.9) | 8.1 (7.3) |

**Table 1.** Effectiveness of single measures: Mean cluster size including 75$^{th}$ and 90$^{th}$ percentile values and mean reproduction number $R$ including standard deviation. For every measure and every school type, values are reported for a student and a teacher source case, respectively. Values are calculated from 500 runs for every (school type, measure, source case) combination.

| type | measure | cluster size, mean (75th,90th percentile) student | teacher | $R$, mean (SD) student | teacher |
|---|---|---|---|---|---|
| primary | No NPIs | 8 (20, 36) | 12 (28, 42) | 1.8 (1.7) | 3.4 (3.0) |
| | + Ventilation | 2 (4, 6) | 3 (6, 10) | 0.9 (0.9) | 1.3 (1.5) |
| | + Masks Teacher | 2 (4, 6) | 2 (2, 6) | 0.8 (0.9) | 0.7 (1.0) |
| | + Masks Students | 1 (2, 4) | 2 (2, 4) | 0.6 (0.7) | 0.6 (0.9) |
| | + Class Size Reduction | 1 (2, 2) | 1 (0, 2) | 0.4 (0.6) | 0.4 (0.7) |
| | Ventilation + Testing Teacher 1x | 2 (4, 6) | 2 (4, 8) | 0.8 (1.0) | 0.9 (1.4) |
| | + Testing Students 1x | 1 (2, 4) | 2 (4, 6) | 0.5 (0.9) | 0.9 (1.3) |
| | + Testing Teacher 2x | 1 (2, 4) | 2 (2, 6) | 0.6 (0.9) | 0.7 (1.2) |
| | + Testing Students 2x | 1 (2, 4) | 2 (2, 6) | 0.4 (0.8) | 0.8 (1.2) |
| | Ventilation, Masks, Testing 1x | 1 (2, 2) | 1 (0, 2) | 0.3 (0.6) | 0.4 (0.7) |
| | Ventilation, Reduction, Testing 1x | 1 (2, 2) | 2 (2, 4) | 0.3 (0.6) | 0.6 (1.0) |
| | All NPIs | 1 (2, 2) | 1 (0, 0) | 0.2 (0.4) | 0.2 (0.5) |
| + day care | No NPIs | 23 (73, 116) | 27 (82, 124) | 2.2 (2.0) | 3.2 (3.0) |
| | + Ventilation | 3 (6, 10) | 4 (6, 14) | 1.0 (1.2) | 1.3 (1.5) |
| | + Masks Teacher | 2 (4, 8) | 2 (2, 6) | 1.0 (1.1) | 0.7 (0.9) |
| | + Masks Students | 1 (2, 4) | 1 (2, 4) | 0.6 (0.8) | 0.5 (0.8) |
| | + Class Size Reduction | 1 (2, 2) | 1 (0, 2) | 0.4 (0.6) | 0.4 (0.7) |
| | Ventilation + Testing Teacher 1x | 2 (4, 10) | 2 (4, 8) | 1.0 (1.1) | 0.9 (1.3) |
| | + Testing Students 1x | 2 (4, 6) | 2 (2, 8) | 0.7 (1.0) | 0.9 (1.4) |
| | + Testing Teacher 2x | 2 (4, 6) | 2 (2, 6) | 0.6 (1.0) | 0.7 (1.2) |
| | + Testing Students 2x | 1 (2, 4) | 2 (2, 4) | 0.5 (0.8) | 0.7 (1.2) |
| | Ventilation, Masks, Testing 1x | 1 (2, 4) | 1 (0, 2) | 0.3 (0.7) | 0.4 (0.7) |
| | Ventilation, Reduction, Testing 1x | 1 (2, 4) | 2 (0, 2) | 0.3 (0.6) | 0.5 (0.9) |
| | All NPIs | 1 (2, 2) | 1 (0, 0) | 0.2 (0.5) | 0.2 (0.6) |
| lower secondary | No NPIs | 57 (172, 192) | 66 (174, 196) | 2.6 (2.1) | 6.5 (5.5) |
| | + Ventilation | 5 (8, 22) | 8 (16, 34) | 1.2 (1.2) | 2.6 (2.5) |
| | + Masks Teacher | 3 (6, 10) | 3 (6, 12) | 1.1 (1.1) | 1.4 (1.6) |
| | + Masks Students | 2 (2, 4) | 2 (4, 6) | 0.7 (0.9) | 1.1 (1.2) |
| | + Class Size Reduction | 1 (2, 2) | 2 (2, 4) | 0.4 (0.7) | 0.7 (1.0) |
| | Ventilation + Testing Teacher 1x | 4 (6, 16) | 5 (8, 18) | 1.2 (1.3) | 1.8 (2.4) |
| | + Testing Students 1x | 2 (4, 8) | 5 (10, 18) | 0.7 (1.1) | 1.7 (2.4) |
| | + Testing Teacher 2x | 2 (2, 8) | 3 (6, 12) | 0.7 (1.1) | 1.4 (2.1) |
| | + Testing Students 2x | 2 (2, 4) | 3 (6, 12) | 0.5 (0.8) | 1.4 (2.1) |
| | Ventilation, Masks, Testing 1x | 1 (2, 2) | 2 (2, 4) | 0.3 (0.7) | 0.6 (1.0) |
| | Ventilation, Reduction, Testing 1x | 1 (2, 4) | 2 (4, 6) | 0.4 (0.7) | 1.1 (1.6) |
| | All NPIs | 1 (2, 2) | 1 (0, 2) | 0.2 (0.5) | 0.4 (0.7) |
| + day care | No NPIs | 49 (148, 176) | 55 (154, 178) | 2.7 (2.3) | 4.3 (3.8) |
| | + Ventilation | 5 (8, 18) | 5 (10, 22) | 1.3 (1.3) | 1.8 (1.9) |
| | + Masks Teacher | 3 (6, 12) | 3 (4, 8) | 1.2 (1.3) | 1.0 (1.3) |
| | + Masks Students | 2 (4, 6) | 2 (2, 4) | 0.7 (1.0) | 0.8 (1.1) |
| | + Class Size Reduction | 1 (2, 2) | 1 (0, 2) | 0.5 (0.7) | 0.5 (0.8) |
| | Ventilation + Testing Teacher 1x | 3 (6, 12) | 4 (4, 14) | 1.2 (1.3) | 1.2 (1.7) |
| | + Testing Students 1x | 2 (4, 8) | 3 (4, 12) | 0.8 (1.2) | 1.1 (1.7) |
| | + Testing Teacher 2x | 2 (4, 8) | 3 (2, 8) | 0.8 (1.2) | 0.9 (1.5) |
| | + Testing Students 2x | 2 (2, 6) | 2 (2, 8) | 0.6 (1.0) | 0.9 (1.4) |
| | Ventilation, Masks, Testing 1x | 1 (2, 4) | 1 (0, 2) | 0.4 (0.7) | 0.4 (0.8) |
| | Ventilation, Reduction, Testing 1x | 1 (2, 4) | 2 (2, 4) | 0.4 (0.7) | 0.8 (1.2) |
| | All NPIs | 1 (2, 2) | 1 (0, 0) | 0.2 (0.5) | 0.3 (0.6) |
| upper secondary | No NPIs | 113 (302, 330) | 122 (306, 332) | 3.4 (2.6) | 7.1 (6.0) |
| | + Ventilation | 8 (16, 34) | 11 (24, 52) | 1.6 (1.5) | 2.8 (2.6) |
| | + Masks Teacher | 4 (10, 16) | 4 (8, 14) | 1.5 (1.4) | 1.4 (1.5) |
| | + Masks Students | 2 (4, 6) | 2 (4, 8) | 0.9 (1.0) | 1.1 (1.2) |
| | + Class Size Reduction | 1 (2, 4) | 2 (2, 4) | 0.6 (0.8) | 0.7 (0.9) |
| | Ventilation + Testing Teacher 1x | 6 (10, 22) | 7 (12, 34) | 1.5 (1.4) | 1.9 (2.5) |
| | + Testing Students 1x | 3 (4, 12) | 6 (12, 28) | 0.9 (1.3) | 1.9 (2.6) |
| | + Testing Teacher 2x | 3 (4, 10) | 4 (6, 16) | 0.8 (1.2) | 1.5 (2.2) |
| | + Testing Students 2x | 2 (2, 6) | 4 (6, 14) | 0.6 (1.1) | 1.4 (2.3) |
| | Ventilation, Masks, Testing 1x | 1 (2, 4) | 2 (2, 4) | 0.5 (0.8) | 0.7 (1.2) |
| | Ventilation, Reduction, Testing 1x | 1 (2, 4) | 3 (4, 8) | 0.5 (0.8) | 1.0 (1.5) |
| | All NPIs | 1 (2, 2) | 1 (0, 2) | 0.2 (0.5) | 0.4 (0.8) |
| secondary | No NPIs | 282 (746, 806) | 306 (760, 826) | 2.9 (2.4) | 8.6 (7.9) |
| | + Ventilation | 9 (13, 42) | 16 (32, 66) | 1.3 (1.3) | 3.3 (3.3) |
| | + Masks Teacher | 4 (8, 16) | 5 (8, 20) | 1.2 (1.2) | 1.6 (1.8) |
| | + Masks Students | 2 (4, 6) | 3 (4, 8) | 0.8 (0.9) | 1.2 (1.5) |
| | + Class Size Reduction | 1 (2, 4) | 2 (2, 4) | 0.5 (0.7) | 0.8 (1.1) |
| | Ventilation + Testing Teacher 1x | 7 (8, 26) | 9 (13, 34) | 1.3 (1.3) | 2.2 (2.9) |
| | + Testing Students 1x | 3 (4, 12) | 9 (16, 38) | 0.8 (1.2) | 2.5 (3.6) |

Continued on next page

| type | measure | cluster size, mean (75th,90th percentile) | | $R$, mean (SD) | |
|---|---|---|---|---|---|
| | | student | teacher | student | teacher |
| | + Testing Teacher 2x | 3 (4, 14) | 7 (10, 28) | 0.9 (1.4) | 1.9 (2.8) |
| | + Testing Students 2x | 3 (4, 6) | 6 (8, 26) | 0.6 (0.9) | 2.0 (2.9) |
| | Ventilation, Masks, Testing 1x | 1 (2, 4) | 2 (2, 6) | 0.4 (0.8) | 0.9 (1.5) |
| | Ventilation, Reduction, Testing 1x | 1 (2, 4) | 4 (4, 12) | 0.5 (0.7) | 1.5 (2.2) |
| | All NPIs | 1 (2, 2) | 2 (0, 2) | 0.3 (0.6) | 0.5 (0.9) |

**Table 2.** Effectiveness of measure combinations: Mean cluster size including 75th and 90th percentile values and mean reproduction number $R$ including standard deviation. For every measure combination and every school type, values are reported for a student and a teacher source case, respectively. Measures preceded by a "+" are added on top of the preceding measures. Values are calculated from 500 runs for every (school type, measure, source case) combination.

#### Supplementary Note 3: Sensitivity analysis

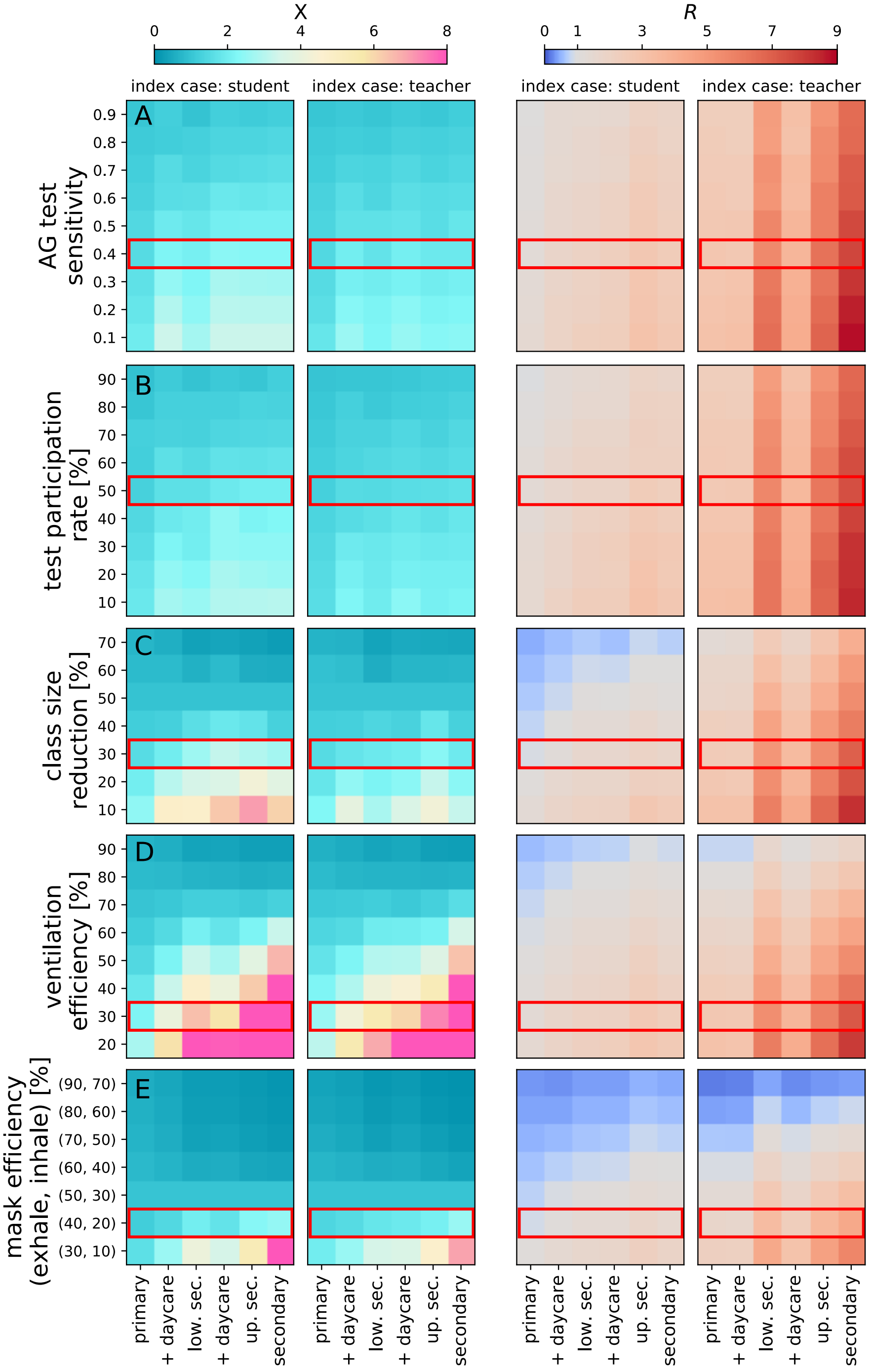


**Fig. 1.** Sensitivity analysis of the efficiency of intervention measures. The two columns to the left show the fold increase of the average cluster size $X$ with respect to the results shown in the main manuscript, Figure **4** (optimistic assumption) for a student and teacher source case, respectively. The two columns to the right show the average number of transmissions from the source case $R$ for a student and teacher source case, respectively. **A**: sensitivity of antigen tests is varied between 0.1 and 0.9 (optimistic assumption: 1.0). **B**: voluntary test participation rate of teachers and students is varied between 10% and 90% (optimistic assumption: 100%). **C**: class size reduction is varied between 10% and 70% (optimistic assumption: 50%). **D**: ventilation efficiency is varied between 20% and 90% (optimistic assumption: 64%). **E**: mask efficiency for exhaling and inhaling is varied between (30%, 10%) and (90%, 70%), respectively (optimistic assumption: (50%, 30%)). For each investigated intervention measure, the conservative assumption shown in Figure **5** (main manuscript) is indicated by a red box.

#### Supplementary Note 4: School questionnaire

*Disclaimer: If you do not tell us otherwise, we will treat all information you give us as confidential. We will not disclose your name or your school's name. In any publication where we use this information, we will just refer to "experts and practitioners from schools". Please inform us if you would like to be acknowledged though.*

*If you do not feel comfortable to answer a question or do not have the information to answer it, feel free to skip it!*

**A. General questions about your school.** *In this section we try to understand how your school is structured and organised. The aim is to understand what kind of contacts occur between different groups of people (teachers, students of the same class, students of different classes) in your school. This is important because during each contact there is the potential to transmit an infection.*

***A.1. Students.***

- Which age groups are taught in your school?
- How many students does your school approximately have?
- Do siblings of students of your school usually also go to your school?
- Does your school have fixed student groups (classes) that stay constant throughout the year and are taught together the majority of the time?
- If no: please describe how the teaching is organised in your school and ignore questions 6 to 13. For example: are students organised in some sort of learning groups? Do students of different age groups mix?
- How many classes does your school have?
- How many students are, on average, in one class?
- Are classes distributed over several floors?
- If yes: are classes with students of the same age group usually located on the same floor?
- Are students sometimes taught together with students of other classes but the **same age group**? If yes: can you estimate how often this is the case? (for example, approximately 1/3 of the lessons).
- Are students sometimes taught together with students of other classes and **different age groups**? If yes: can you estimate how often this is the case? (for example, approximately 1/3 of the lessons).
- Is the assumption correct that students usually have the large majority of their friends within the same class?
- How many friends (on average) do students have in classes other than their own? *We know that this is a very hard question to answer and we would already be very happy if you can give us a very rough estimate!*
- Are there shared areas in which students of different classes meet during breaks, for example a courtyard or canteen?
- Is there other information that you think would be important for us to understand how teaching is organised and how transmissions of an infection could occur at your school?

***A.2. Teachers.***

- How many teachers are working at your school?
- If your school does have classes: How many different classes, on average, does a teacher teach
  - a Over the course of a week?
  - b Over the course of a day?
  - c Did this change between now and the time before the second lockdown (weeks 36-45)?
- Is the assumption correct that a lesson is usually supervised by a single teacher?
- Are there areas in which teachers regularly meet (faculty room, canteen)?
- Is the assumption correct that every teacher meets the majority of their colleagues at least once a day, for example during lunch? If no: please describe which teachers usually meet at your school (for example: teachers teaching the same subject meet more often, all teachers meet only once a week, etc.).

**B. Questions regarding strategies to prevent the spread of COVID-19.** *In this section we try to understand which measures your school has implemented to prevent the spread of COVID-19, and how these measures influence the organisation of teaching. In addition, it is very important for us to understand how testing and isolation of (potentially) positive cases at your school works.*

##### *B.1. Organisation of teaching.*

- Masks for teachers:
  - a Do teachers wear masks during lessons?
  - b Did teachers wear masks during lessons before the second lockdown (weeks 36-45)?
  - c Are teachers recommended to wear masks if they meet other teachers?
- Masks for students:
  - a Do students wear masks during lessons? Is this different for students from different age groups?
  - b Did students wear masks during lessons before the second lockdown (weeks 36-45)?
  - c Do students wear masks outside of lessons when they move through the building?
- Size of classes:
  - a Is the number of students that participate in in-person teaching somehow reduced (for example halving of classes)?
  - b Was this already the case before the second lockdown (weeks 36-45)?
- Are students discouraged from meeting students from other classes on the hallways between lessons?
- Are there measures in place that prevent the mixing of students from different classes in the canteen and/or courtyard during longer breaks?
- Are there measures that regulate which teachers meet which other teachers? (For example only teachers that teach the same classes are allowed to meet).
- Did the way your school organises in-person teaching change between now and before the second lockdown (weeks 36-45) regarding
  - a A change on how classes are distributed in the building?
  - b A change in the number of different classes that are taught by a teacher?
  - c A reduction of the number of lessons in which students from different classes are taught together?
  - d A reduction of in-person teaching (in favour of more online teaching)?
  - e A reduction or cancellation of high-risk courses such as singing or sports?
  - f Other changes?
- Do you think the implementation of the following recommendations works reasonably well at your school:
  - a Frequent washing of hands?
  - b Frequent airing of rooms?
  - c Wearing of masks?
- Are there additional preventive measures implemented at your school that you think would be important for us to know?

##### *B.2. Testing and quarantine.*

- Is there any regular testing procedure in place at your school for
  - a All teachers? If yes: in which intervals?
  - b All students? If yes: in which intervals?
  - c Was this already the case before the second lockdown (weeks 36-45)?
- If a student **at school** is suspected of being infected with COVID-19:

a Is the student tested at the school?
b Is the student sent home?
c How long does it usually take until there is a test result?
d Which precautions are taken for students that were in contact with the suspected student?
e Which precautions are taken for teachers that were in contact with the suspected student?

- If a student **at home** is suspected of being infected with COVID-19:

a Is the student isolated until they have a negative test result (i.e. not allowed to go to school)?
b How long does it usually take until there is a test result?
c Are any siblings of that student that go to the same school also asked to stay at home?
d Which precautions are taken for students that were in contact with the suspected student?
e Which precautions are taken for teachers that were in contact with the suspected student?

- If a student has a positive test result:

a Are any classmates of the student isolated?
b If yes: is the whole class isolated or only selected students? How is it decided which students are isolated?
c For how long are students usually isolated?
d Are any contact persons of the positive student tested?

- Have there been any changes regarding the testing and isolation of students between now and the time before the second lockdown (weeks 36-45)?

- If a teacher **at school** is suspected of being infected with COVID-19:

a Is the teacher tested at school?
b Is the teacher required to go home?
c How long does it usually take until there is a test result?
d Which precautions are taken for students that were in contact with the suspected teacher?
e Which precautions are taken for teachers that were in contact with the suspected teacher?

- If a teacher **at home** is suspected of being infected with COVID-19:

a Is the teacher isolated until the test result arrives (i.e. not allowed to come to work)?
b How long does it usually take until there is a test result?
c Which precautions are taken for students that were in contact with the suspected teacher?
d Which precautions are taken for teachers that were in contact with the suspected teacher?

- If a teacher has a positive test result:

a Are other teachers that had a high-risk contact with the positive teacher isolated?
b If yes: How do you determine what qualifies as a high-risk contact?
c For how long are teachers usually isolated?
d Are any students that were taught by the teacher isolated?
e Are any contact-persons of the positive teacher tested?

- Have there been any changes regarding the testing and isolation of teachers between now and the time before the second lockdown (weeks 36-45)?

- Is there any additional information that you think would be important for us to understand how your school handles (potentially) positive COVID-19 cases?

*We would like to thank you for taking your valuable time to answer our questions! The information you give us will provide us with valuable insights to inform our model of the spread of the corona virus in Austrian schools and help to make the model as close to reality as possible.*

### Bibliography


1. Science Federal Ministry for Education and Republic of Austria Research. Grunddaten des österreichischen schulwesens. Technical report, Federal Ministry for Education, Science and Research, Republic of Austria, 2018.
2. Karoline Mitterer and Marion Seisenbacher. Fact sheets: Pflichtschule und tagesbetreuung, 2020.
3. Science Federal Ministry for Education and Republic of Austria Research. Bedarfsgerechter ausbau der ganztägigen schulformen / standorte 2020/21. Technical report, Federal Ministry for Education, Science and Research, Republic of Austria, 2020.
4. Science Federal Ministry for Education and Republic of Austria Research. Lehrplan der volksschule, vierter teil, gesamtstundenzahl und stundenausmaß der pflichtgegenstände, der verbindlichen Übungen, des förderunterrichts, der freigegenstände und unverbindlichen Übungen. Technical report, Federal Ministry for Education, Science and Research, Republic of Austria, 2005.
5. Sonderschule und inklusiver unterricht. https://www.bmbwf.gv.at/Themen/schule/schulsystem/sa/sp.html, 2021. Accessed: 2021-01-29.
6. Mittelschule. https://www.bmbwf.gv.at/Themen/schule/schulsystem/sa/ms.html, 2021. Accessed: 2021-01-29.
7. Gesamte rechtsvorschrift für lehrpläne der mittelschulen. https://www.ris.bka.gv.at/GeltendeFassung.wxe?Abfrage=Bundesnormen&Gesetzesnummer=20007850, 2021. Accessed: 2021-01-29.
8. Lehrplan der allgemeinen sonderschule. https://www.ris.bka.gv.at/Dokumente/BgblAuth/BGBLA_2008_II_137/COO_2026_100_2_440355.html, 2021. Accessed: 2021-01-29.
9. Allgemeinbildende höhere schulen. https://www.oesterreich.gv.at/themen/bildung_und_neue_medien/schule/2/Seite.1760160.html, 2021. Accessed: 2021-01-29.
10. Berufsbildende mittlere und höhere schulen. https://www.bmbwf.gv.at/Themen/schule/schulsystem/sa/bmhs.html, 2021. Accessed: 2021-01-29.
11. Gesamte rechtsvorschrift für lehrpläne – allgemeinbildende höhere schule. https://www.ris.bka.gv.at/GeltendeFassung.wxe?Abfrage=Bundesnormen&Gesetzesnummer=10008568, 2021. Accessed: 2021-01-29.
